# Temporal trends of dengue seroprevalence among children in coastal Kenya, 1998–2018: a longitudinal cohort study

**DOI:** 10.1101/2023.08.13.23294039

**Authors:** Henry K. Karanja, Benedict Orindi, John N. Gitonga, Daisy Mugo, Kennedy Mwai, Doris K. Nyamwaya, Donwilliams Omuoyo, Barnes S. Kitsao, Jennifer N. Musyoki, Julianna Wambua, Edward Otieno, Lynette Isabella Ochola-Oyier, Philip Bejon, George M. Warimwe

## Abstract

Serosurveys suggest widespread dengue virus (DENV) transmission in Africa but there is limited information on the temporal patterns of exposure. Here, we estimated the prevalence and incidence of DENV infections in coastal Kenya over a 20-year period (1998–2018). Sera (n=8038) obtained annually from a longitudinal cohort of 1847 children aged 15 years and below were screened for anti-DENV IgG antibodies. Anti-DENV IgG seroprevalence increased with age and peaked during outbreak years. Among 1354 children who were seronegative at recruitment, we observed an overall incidence (seroconversion) rate of 129.5 (95% CI 118.7–141.4) DENV infections per 1000 person-years. The highest incidence was observed in 2013 at 520 infections per 1000 person-years (95% CI 443.6–610.2) coinciding with a large DENV outbreak in coastal Kenya. Our data suggest long-term DENV exposure among children in coastal Kenya highlighting an urgent need for clinical surveillance to determine the associated health burden in this setting.

## Introduction

Dengue virus (DENV) is the most widespread arbovirus globally, causing over 300 million infections every year (*1*). This positive-stranded RNA virus belongs to the genus flavivirus and is composed of a genome, approximately 11kb in size, that exists in four distinct serotypes (DENV-1 to DENV-4) that can be further classified into multiple genotypes (*2*). Infection results in long-lived protection against the infecting DENV serotype but only short-lived cross-protection against heterologous serotypes (*3*). DENV is primarily transmitted by *Aedes aegypti* mosquitoes whose wide geographic distribution accounts for the high global burden of the disease (*4, 5*). Most DENV infections are asymptomatic, but an estimated 100 million clinical cases occur every year with manifestations ranging from a mild self-limiting febrile illness to life-threatening symptoms such as hemorrhagic fever and shock syndrome (*2*). Licensed dengue vaccines are now available for use in settings where the disease in endemic (*6, 7*). Knowledge of the burden and geographic distribution of DENV infections is essential for efficient deployment and use of dengue vaccines and other disease control interventions.

The burden of DENV infections in Africa is poorly defined with most literature focusing on disease outbreaks in Asia and the Americas (*8*). A modelling study suggests that Africa accounts for more than 16% of the global burden of DENV infections annually (*1*). The limited seroprevalence and outbreak report data available support widespread viral transmission on the continent (*9*), but there are insufficient data on the age-specific risk of DENV exposure to inform local prioritization for clinical surveillance and public health response. In Kenya, dengue outbreaks have been reported since 1982 with wide geographic risk of exposure (*10-12*). DENV-2 was implicated in the initial 1982 outbreak in coastal Kenya (*10*), and though other serotypes are in circulation (*11-13*), DENV-2 is most frequently associated with outbreaks in Kenya and other settings in Africa (*9, 14*). For instance, recent clinical surveillance in western and coastal Kenya during 2014 to 2017 identified a high prevalence of DENV viraemia among children with undifferentiated fever (42% of 862 patients); while viral genomes from all 4 DENV serotypes were detected, DENV-2 accounted for more than half of the sequenced genomes (*13*). Together, these data suggest high levels of DENV exposure in Kenya, but the extent to which exposure has varied over time remains unknown.

Here, we use a biobank to investigate the temporal trends of DENV exposure in coastal Kenya where the bulk of DENV outbreaks in Kenya have been reported. We used immunoglobin G (IgG) serology to estimate the age-specific prevalence of anti-DENV IgG antibody among children aged between 6 months and 15 years over a 20-year period, that is, 1998 to 2018.

## Materials and methods

### Study population

This study was nested within an ongoing longitudinal cohort study at the Kenya Medical Research Institute (KEMRI)-Wellcome Trust Research Programme in Kilifi, coastal Kenya conducted since 1998 to date with the main aim of describing malaria incidence and transmission in this area (*15, 16*). Briefly, children resident in Junju and Ngerenya sub-locations within the Kilifi Health and Demographic Surveillance System (KHDSS) (*17*) are recruited at birth and remain in the cohort until the age of 15 years. The malaria surveillance has been running in Ngerenya since 1998 and in Junju since 2005. Every year, a cross-sectional survey is conducted and includes blood sampling of cohort participants and collection of demographic data. Sera (and linked demographic data) collected during these annual cross-sectional surveys were available at the KEMRI-Wellcome Trust Research Programme biobank, allowing an assessment of DENV seroprevalence over time. Ethical approval was provided by the KEMRI Scientific and Ethics Review Unit (KEMRI SERU # 3296, #1131).

### Estimation of DENV exposure by serology

We measured anti-DENV IgG antibodies by enzyme-linked immunosorbent assay (ELISA) against a local DENV-2 isolate obtained from a patient in the same geographic setting (*18*). DENV-2 was selected for this assay owing to its frequent circulation in Kenya and ready availability of DENV-2 virus in our lab (*13, 19, 20*). Briefly, 96 wells Maxisorp flat-bottomed plates were coated with 50μL of ∽1×10^3^ TCID_50_ whole DENV-2 virus lysate per well and incubated for 1 hour at room temperature (RT), fixed with 4% paraformaldehyde for 30 minutes at RT and blocked with Blocker™ Casein in Phosphate Buffered Saline (PBS) (ThermoScientific, cat. # 37528). Serum samples were diluted 1:500 in Blocker™ Casein and added to the DENV-coated plates and incubated for 2 hours at 37°C. The plates were washed 3 times with wash buffer (0.1% Tween20 in PBS), after which 1:10000 dilution of Peroxidase Labelled Goat Anti-Human IgG (Cat. # 074-1002, KPL) in wash buffer was added to the plates and incubated for one hour. After another series of washes, the plates were developed by adding 100μL/well of reconstituted o-phenylenediamine dihydrochloride (Sigma, cat. # 8287) for 15–20 minutes and the reaction stopped by adding 100μl/well of 2NH_2_SO_4_. Colour reaction was measured at 492nm by the Tecan Infinite® 200 PRO. Each plate included serial dilutions of a pooled sera sample from six Kenyan adults with high virus neutralising antibody titres against DENV-2 from which a standard curve was generated and used to extrapolate IgG responses (expressed as ELISA units) for each test sample. To define a seropositivity cut-off we used the whole virus IgG ELISA to test sera from 108 adults with corresponding focus reduction neutralisation test (FRNT_90_) data (*18*) and determined the sensitivity and specificity of the IgG ELISA in detecting DENV-2 FRNT_90_ positive sera. An IgG ELISA unit cut-off ≥250 had a specificity of 91.1% and sensitivity of 92.1% in identifying individuals with DENV-2 neutralising antibodies as measured by FRNT_90_ assay and was therefore used for the analysis presented herein. Because IgG ELISA cannot reliably distinguish DENV serotype-specific responses (*2*), which requires use of the highly specific virus neutralisation assays, we describe the IgG immune responses as anti-DENV rather than anti-DENV-2.

## Statistical analysis

First, we estimated DENV seroprevalence, together with the associated 95% Agresti-Coull confidence interval by sex, age, location of residence (Junju or Ngerenya) and year of sampling. Next, to assess whether the seroprevalence varied with sex, age, location and year we fitted a mixed-effects logistic regression model, with child as a random effect. Age entered the model as a linear spline function with a single knot at 10 years (*21*), such that we estimated the linear effect of age up to the age of 10 years then the linear effect of age after 10 years. The covariates were tested using the likelihood ratio (LR) test. Finally, we estimated incidence rates (IR) of DENV infection (defined as IgG seropositivity in the whole virus ELISA) among children and compared across variables using Poisson regression, with the logarithm of person-years as offset. This analysis only included children who were initially DENV seronegative at recruitment, and children were censored after their first DENV infection (that is, when they turned IgG seropositive). We report incidence rates per 1000 person-years. Data were analysed using Stata version 17.0 (StataCorp, College Station, TX) and graphs generated using GraphPad Prism version 9.5.1 (GraphPad Software, San Diego, California USA). All tests were performed at 0.05 significance level.

## Results

A total of 8038 serum samples obtained from 1847 children at annual cross-sectional surveys between 31^st^ August 1998 and 10^th^ April 2018 were available in the KEMRI-Wellcome Trust Research Programme biobank. The median number of annual cross-sectional surveys (or years of follow-up) for the 1847 children was 2 (range 1–14), contributing to a total of 7394.2 years of follow-up over the 20-year period. The median age of the children over the entire follow-up period was 7.1 years (interquartile range 4.1–10.3). Of the 8038 serum samples 3965 (49.3%) came from children resident in Junju and 4073 (50.7%) were from residents of Ngerenya (Table 1).

**Table 1.** Prevalence and incidence rates of DENV infection among children in Kilifi by sex, age and location.

|  | Number of children | Number of samples | Samples positive | Seroprevalence |  |  | Person years* | Incidence |  |  |  |
| --- | --- | --- | --- | --- | --- | --- | --- | --- | --- | --- | --- |
|  |  |  |  | % positive (95% CI) | Adjusted odds ratio (95% CI) | P-value |  | Cases | Incidence rate/1000 person-years (95% CI) | Incidence rate ratio (95% CI) | P-value |
| Overall | 1847 | 8038 | 2295 | 28.6 (27.6–29.5) |  |  | 3893.2 | 503 | 129.2 (118.4–141.0) |  |  |
| Sex |  |  |  |  |  |  |  |  |  |  |  |
| Female | 900 | 3849 | 1089 | 28.3 (26.9–29.7) | Reference |  | 1875.2 | 239 | 127.5 (112.3–144.7) | Reference |  |
| Male | 947 | 4189 | 1206 | 28.8 (27.4–30.2) | 1.10 (0.87–1.38) | 0.438 | 2018.0 | 264 | 130.8 (116.0–147.6) | 1.02 (0.85–1.21) | 0.841 |
| Age in years** |  |  |  |  |  |  |  |  |  |  |  |
| Age spline 1 |  |  |  |  | 1.46 (1.40–1.52) | <0.001 |  |  |  | 1.05 (1.01–1.10) | 0.019 |
| Age spline 2 |  |  |  |  | 0.97 (0.92–1.03) | 0.310 |  |  |  | 0.98 (0.90–1.106) | 0.564 |
| Location |  |  |  |  |  |  |  |  |  |  |  |
| Junju | 907 | 3965 | 1019 | 25.7 (24.4–27.1) | Reference |  | 1680.9 | 249 | 148.1 (130.8–167.7) | Reference |  |
| Ngerenya | 940 | 4073 | 1276 | 31.3 (29.9–32.8) | 0.97 (0.73–1.29) | 0.834 | 2212.2 | 254 | 114.8 (101.5–129.8) | 0.91 (0.74–1.12) | 0.365 |

\*Person-years contributed by the 1847 children up to the point of their first DENV infection detection, beyond which children were censored.
\*\*Non-linear age effect was modelled using a 1-knot linear spline. Age spline 1 represent the continuous age measurements in the range 0.5 to 10; age spline 2 represent the
continuous age measurements in the range 10–15 years. Age splines 1 and 2 estimate the linear effect of age in those two age ranges.

Overall, 2295 (28.6%, 95% CI 27.6–29.5%) of the 8038 samples were seropositive for DENV IgG antibodies. However, DENV seroprevalence markedly increased with age from a low of 2.2% among children aged below 1 year to a high of 40% among those aged 9 and 10 years (Odds ratio (OR) 1.46, 95% CI 1.40–1.52; p<0.001), after which the seroprevalence remained the same (OR 0.97, 95% CI 0.92–1.03) (Figure 1C and Table 1). DENV seroprevalence varied substantially over the years (LR test p<0.0001) with seroprevalence as high as 60% observed in 1998 and 2013, to a low of approximately 12% in 2017 (Figure 1A) but did not vary significantly by sex (p=0.438) and location (p=0.834) (Table 1).

**Figure 1.**
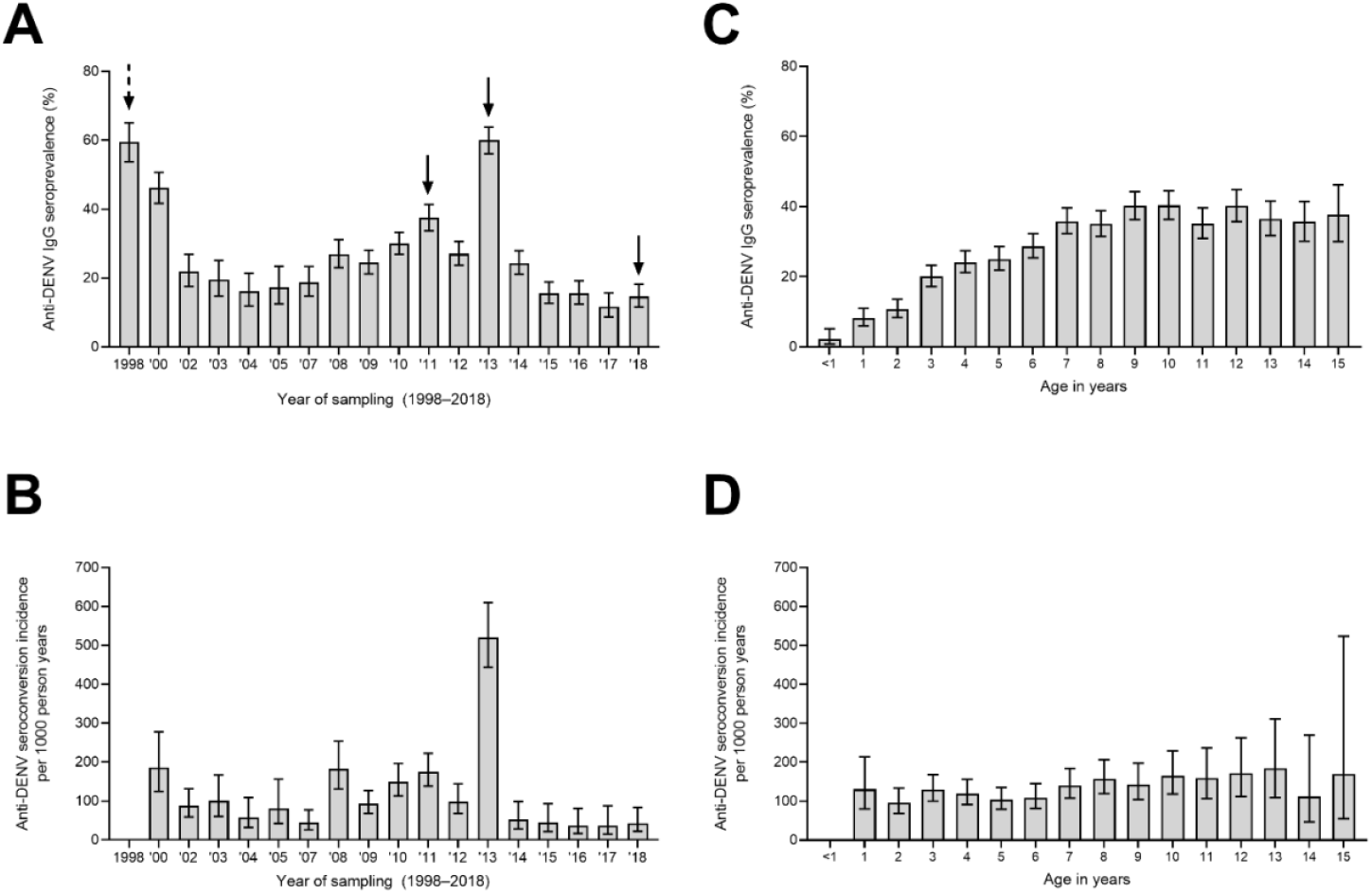
Prevalence and incidence of DENV IgG antibodies by year of sampling and age of the children. Presented in (A) are the percentages of children with anti-DENV IgG antibodies in each annual cross-sectional survey during the study period. In (B) the incidence of anti-DENV IgG seroconversion among children who were seronegative for DENV at recruitment is shown over time. (C) presents the percentages of children with anti-DENV IgG antibodies from 1998–2018 by 1-year age bands. (D) presents the incidence of anti-DENV IgG seroconversion among children who were seronegative for DENV at recruitment by 1-year age bands. An incidence estimate could not be made for the year 1998 and for children aged below 1 year as participants did not have a positive time at risk. The bold arrows indicate when dengue outbreaks occurred in Kenya (solid) and in neighbouring countries (dashed).

During the 20-year period, we observed 503 new DENV infections (i.e., seroconversion among 1354 participants who were seronegative at recruitment), giving an overall incidence rate of 129.2 (95% CI 118.4–141.0) DENV infections per 1000 person-years. The incidence of seroconversion did not vary by sex (incidence rate ratio (IRR) 1.02, 95% CI 0.85–1.21; p=0.841) and geographical location of residence (p=0.365), but it linearly increased with age at the rate of 5% per one year increase in age up to around the age of 9–10 years (IRR 1.05, 95% CI 1.01–1.10; p=0.019) (Table 1 and Figure 1D). The highest incidence of seroconversion was observed in 2013 at 520 cases per 1000 person-years (95% CI 443.6–610.2), followed by 2008 at 182.3 cases per 1000 person-years (95% CI 130.9– 253.9) and 2011 at 174.8 cases per 1000 person-years (95% CI 137.8–221.7) (Figure 1B; LR test p<0.001).

## DISCUSSION

There is limited surveillance for DENV in Africa beyond sporadic reporting of outbreaks (*22*). The true burden of DENV infections therefore remains unknown with modelling studies suggesting a burden in Africa that is at least as high as that in Latin America (*1*). Most of the previous serological assessments of DENV exposure in Africa have tended to be cross-sectional in nature, providing information on the geographical extent of DENV transmission but limited information on the temporal dynamics of viral exposure (*14*). Our data provide a two-decade assessment of DENV infections in coastal Kenya.

We confirm high levels of DENV exposure in coastal Kenya, in keeping with previous serological studies and outbreak reports (*11, 12, 18*). DENV seropositivity showed a positive relationship with age suggesting ongoing endemic transmission in this setting. Seropositivity peaked in 1998–2000 and 2013, coinciding with periods when DENV cases were reported in neighbouring countries (Tanzania and Uganda in 1999–2000 (*14*)) and with one of the largest DENV outbreaks in coastal Kenya reported in 2013 (*23*). During the 2013 outbreak, DENV cases were predominantly reported in Mombasa (*23*). However, the very high incidence of DENV seroconversion observed in Kilifi, approximately 50km north of Mombasa, during this period suggests that the outbreak was more widespread along the Kenyan coast than previously thought (*11*). We also detected high incidence of DENV seroconversion during 2008 and 2011, coinciding with a confirmed DENV outbreak in northern Kenya and neighbouring Somalia in 2011 (*24*), and a possible undetected outbreak in 2008 in coastal Kenya. These incidence data support the utility of serology in inferring exposure to DENV infections.

DENV-2 has been implicated in most outbreaks and case reports in Africa, but other DENV serotypes are increasingly being detected during outbreaks, including importations from Asia (*9, 14*). Increasing circulation of multiple DENV serotypes is of public health concern due to the potential for severe disease associated with serial infection with heterologous DENV serotypes (*25*). Two types of immune responses are elicited by infection: a long-lived protective antibody response against the infecting DENV serotype and a short-lived cross-reactive response against other (heterologous) DENV serotypes (*2, 3*). During subsequent infection(s) with a heterologous DENV serotype, these cross-reactive weaker antibody responses are unable to neutralize virus infection but instead facilitate Fc-mediated virus entry into host cells (e.g. monocytes) allowing the virus to thrive whilst evading other host anti-viral defenses, resulting in enhanced pathology that underlies severity disease (*2, 25*). Our assay could not distinguish DENV serotype-specific responses. However, the high levels of DENV exposure observed indicate an urgent need for long-term systematic clinical and immunological surveillance to characterise DENV infections in this setting, the associated symptoms and outcomes, and the predisposing risk factors. All four DENV serotypes have been associated with febrile illness in children in Kenya (*13*), but interestingly, the current WHO clinical diagnostic criteria for dengue performed poorly in this setting with no severe complications observed (*26*). This further highlights a need for development of clinical algorithms that can identify patients with DENV infection in East Africa, both for clinical diagnosis and for use in evaluation of disease control interventions.

This study had limitations. First, we used whole DENV-2 virus for the detection of circulating IgG antibodies and while assay performance was good in comparison to virus neutralising responses, the assay cannot distinguish DENV serotype-specific responses and it is likely there were cross-reactive responses against other DENV serotypes and related flaviviruses (e.g., Zika virus). The relative contribution of the four DENV serotypes to the observed exposure levels could therefore not be determined. Nevertheless, we could detect incident cases during periods when DENV outbreaks were reported in Kenya (*23*), and our previous work suggests limited circulation of Zika virus in the study location (*18*). Second, this study provides an unprecedented 20-year longitudinal assessment of DENV exposure in coastal Kenya. While this has allowed a long-term view of exposure trends, the limited geographical focus precludes generalisation to other settings in the country and the region. However, previous dengue outbreaks have tended to involve multiple countries in East Africa, and the shared ecology supportive of the *Aedes spp* and other mosquito vectors suggests DENV infections (as with other arboviruses) may be widespread in the region (*8, 27, 28*). Finally, we did not monitor clinical illness and therefore could not determine what fraction of the observed exposures had experienced symptomatic infections. With licensed dengue vaccines now available (*6, 7*), an assessment of the disease burden in Kenya and other African countries is urgently needed to inform decisions on vaccine deployment and use.

## Data Availability

All data produced in the present work are contained in the manuscript

## Acknowledgements

This work was funded by Wellcome Trust grant (grant number 203077_Z_16_Z to PB) and an Oak Foundation fellowship to GMW. The funders had no role in study design, data collection and analysis, decision to publish, or preparation of the manuscript.

